# Federated Learning of Electronic Health Records Improves Mortality Prediction in Patients Hospitalized with COVID-19

**DOI:** 10.1101/2020.08.11.20172809

**Authors:** Akhil Vaid, Suraj K Jaladanki, Jie Xu, Shelly Teng, Arvind Kumar, Samuel Lee, Sulaiman Somani, Ishan Paranjpe, Jessica K De Freitas, Tingyi Wanyan, Kipp W Johnson, Mesude Bicak, Eyal Klang, Young Joon Kwon, Anthony Costa, Shan Zhao, Riccardo Miotto, Alexander W Charney, Erwin Böttinger, Zahi A Fayad, Girish N Nadkarni, Fei Wang, Benjamin S Glicksberg

## Abstract

Machine learning (ML) models require large datasets which may be siloed across different healthcare institutions. Using federated learning, a ML technique that avoids locally aggregating raw clinical data across multiple institutions, we predict mortality within seven days in hospitalized COVID-19 patients. Patient data was collected from Electronic Health Records (EHRs) from five hospitals within the Mount Sinai Health System (MSHS). Logistic Regression with L1 regularization (LASSO) and Multilayer Perceptron (MLP) models were trained using local data at each site, a pooled model with combined data from all five sites, and a federated model that only shared parameters with a central aggregator. Both the federated LASSO and federated MLP models performed better than their local model counterparts at four hospitals. The federated MLP model also outperformed the federated LASSO model at all hospitals. Federated learning shows promise in COVID-19 EHR data to develop robust predictive models without compromising patient privacy.

## INTRODUCTION

### Background and Significance

Coronavirus disease 2019 (COVID-19) has led to over 700,000 deaths worldwide and other devastating outcomes.[1] Clinical manifestations occur in heterogeneous patterns that are not yet well understood, creating an urgent need to understand factors that lead to negative outcomes to aid delivery of more tailored treatments. This task requires learning from large, diverse patient populations. However, pertinent patient data are siloed within hospitals worldwide. While many studies have produced significant findings in regards to COVID-19 outcomes using data from single hospitals, larger representation from additional populations is needed to derive generalizable outcomes, especially within the context of machine learning (ML).[2-11] Large-scale initiatives such as the National COVID Cohort Collaborative and the 4CE Consortium have joined globally distributed hospitals to examine association analyses, but meta-analysis does not allow joint machine learning from diverse patient data.[12-13]

In light of patient privacy, federated learning is emerging within the biomedical field as a promising strategy particularly in the context of COVID-19 where patient data are fragmented across health systems which are not representative of the entire population, leading to predictions that may not be generalizable.[14] Briefly, federated learning allows for decentralized refinement of independently built ML models. This is done via iterative exchange of model parameters to a central aggregator without sharing raw data.

A limited number of studies have assessed ML models within a federated learning framework for outcome prediction in COVID-19 but show promise. Kumar et al. built a blockchain-based federated learning schema and achieved enhanced sensitivity for COVID-19 detection from lung CT scans.[15] Xu et al. used deep learning to identify COVID-19 on CT scans from multiple hospitals in China [16] and found that models built on hospitals in one region did not generalize well to others, but were able to achieve considerable performance gains when using a federated learning approach. For more detailed background regarding COVID-19, ML in the context of COVID-19, challenges for multi-institutional collaborations, and federated learning, see Supplementary Materials.

While it has been proposed, to our knowledge there are no published studies that implement and assess the utility of federated learning to predict COVID-19 outcomes from Electronic Health Record (EHR) data.[17] In this work, we are the first to build federated learning models to predict mortality in patients diagnosed with COVID-19 within seven days of hospital admission using EHR data.

## MATERIALS AND METHODS

### Clinical Data Source and Study Population

Data from COVID-19 positive patients (n=4029) were derived from Epic EHR systems of five hospitals within the Mount Sinai Health System (MSHS) in New York City (NYC): Mount Sinai Brooklyn (MSB; n=611); Mount Sinai Hospital (MSH; n=1644); Mount Sinai Morningside (MSM); n=749); Mount Sinai Queens (MSQ; n=540); and Mount Sinai West (MSW; n=485). A schematic of all study inclusion criteria is shown in Figure 1A, further detailed in Supplementary Materials. Details on cross-hospital demographic and clinical comparisons are also in the Supplementary Materials.

**Figure 1.**
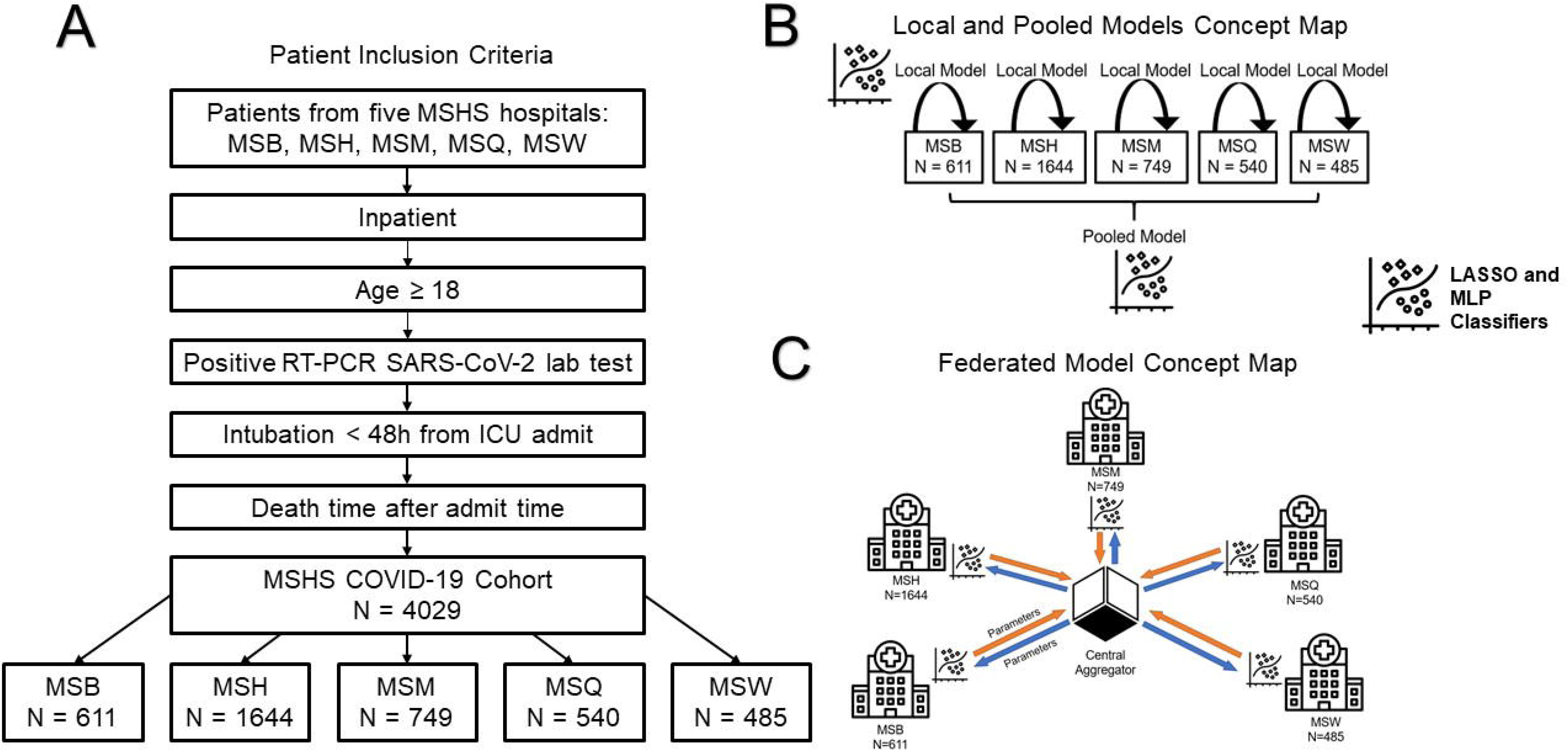
Study Design and Model Workflow. (A) Criteria for patient inclusion in study. (B) Overview of local and pooled models. Local models only utilize data from the site itself while pooled models incorporate data from all sites. Both local and pooled MLP and LASSO models were utilized. (C) Overview of federated model. Parameters from a central aggregator are shared with each site, and sites do not have direct access to clinical data from others. After models are trained locally at a site, parameters with and without added noise are sent back to the central aggregator to update federated model parameters. A federated LASSO and federated MLP model were utilized.

### Study Design

We performed multiple experiments as outlined in Figures 1B and 1C (see Model Development and Selection). First, we developed classifiers using and testing on local data from each hospital separately. We also built a federated learning model by averaging the parameters of models from each individual hospital. Finally, we combined all individual hospital data into a superset to develop a “pooled” model. Although the pooled model is not practically feasible, it represents an ideal framework.

Data included demographics, past medical history, vital signs, lab tests, and outcomes for all patients (Table 1, Supplementary Table 1). Due to varying prevalence across hospitals, we assessed multiple class balancing techniques (Supplementary Table 2). To simulate federated learning in practice, we also performed experiments with the addition of Gaussian noise (see Supplementary Materials).

**Table 1:** Demographics of Hospitalized COVID-19 Patients.

|  | Mount Sinai<br>Brooklyn<br>(MSB) | Mount Sinai<br>Hospital<br>(MSH) | Mount Sinai<br>Morningside<br>(MSM) | Mount Sinai<br>Queens<br>(MSQ) | Mount<br>Sinai West<br>(MSW) | P-Value |
| --- | --- | --- | --- | --- | --- | --- |
| (N) | 611 | 1644 | 749 | 540 | 485 |  |
| Gender, n (%) |  |  |  |  |  |  |
| Male | 338 (55.3) | 951 (57.8) | 411 (54.9) | 344 (63.7) | 257 (53.0) | 0.004 |
| Female | 273 (44.7) | 693 (42.2) | 338 (45.1) | 196 (36.3) | 228 (47.0) |  |
| Age, median (IQR) | 72.5<br>[63.6,82.7] | 63.3<br>[51.3,73.2] | 69.8<br>[57.4,80.3] | 68.1<br>[57.1,78.8] | 66.3<br>[52.5,77.6] | <0.001 |
| Ethnicity, n (%) |  |  |  |  |  |  |
| Hispanic | 21 (3.4) | 460 (28.0) | 259 (34.6) | 198 (36.7) | 111 (22.9) | <0.001 |
| non-Hispanic | 416 (68.1) | 892 (54.3) | 452 (60.3) | 287 (53.1) | 349 (72.0) |  |
| Unknown | 174 (28.5) | 292 (17.8) | 38 (5.1) | 55 (10.2) | 25 (5.2) |  |
| Race, n (%) |  |  |  |  |  |  |
| Asian | 13 (2.1) | 83 (5.0) | 16 (2.1) | 56 (10.4) | 27 (5.6) | <0.001 |
| Black/African American | 323 (52.9) | 388 (23.6) | 266 (35.5) | 64 (11.9) | 109 (22.5) |  |
| Other | 54 (8.8) | 705 (42.9) | 343 (45.8) | 288 (53.3) | 164 (33.8) |  |
| Unknown | 27 (4.4) | 87 (5.3) | 25 (3.3) | 14 (2.6) | 14 (2.9) |  |
| White | 194 (31.8) | 381 (23.2) | 99 (13.2) | 118 (21.9) | 171 (35.3) |  |
| Past Medical History |  |  |  |  |  |  |
| Acute Myocardial Infarction | 14 (2.3) | 16 (1.0) | --- | 15 (2.8) | 7 (1.4) | 0.006 |
| Acute Respiratory Distress Syndrome (ARDS) | --- | 28 (1.7) | --- | --- | --- | <0.001 |
| Acute Venous Thromboembolism | --- | 11 (0.7) | --- | --- | --- | 0.74 |
| Asthma | --- | 100 (6.1) | 39 (5.2) | 19 (3.5) | 27 (5.6) | <0.001 |
| Atrial Fibrillation | 23 (3.8) | 113 (6.9) | 44 (5.9) | 49 (9.1) | 28 (5.8) | 0.005 |
| Cancer | 22 (3.6) | 190 (11.6) | 47 (6.3) | 21 (3.9) | 41 (8.5) | <0.001 |
| Chronic Kidney Disease (CKD) | 46 (7.5) | 208 (12.7) | 75 (10.0) | 81 (15.0) | 33 (6.8) | <0.001 |
| Chronic Obstructive Pulmonary Disease (COPD) | 11 (1.8) | 64 (3.9) | 31 (4.1) | 28 (5.2) | 19 (3.9) | 0.044 |
| Chronic Viral Hepatitis | --- | 17 (1.0) | 14 (1.9) | --- | --- | 0.021 |
| Coronary Artery Disease (CAD) | 56 (9.2) | 168 (10.2) | 92 (12.3) | 82 (15.2) | 51 (10.5) | 0.008 |
| Diabetes mellitus | 93 (15.2) | 351 (21.4) | 165 (22.0) | 154 (28.5) | 76 (15.7) | <0.001 |
| Heart Failure | 36 (5.9) | 110 (6.7) | 61 (8.1) | 43 (8.0) | 30 (6.2) | 0.383 |
| Human Immunodeficiency Virus (HIV) | --- | 32 (1.9) | 11 (1.5) | --- | 14 (2.9) | 0.001 |
| Hypertension | 112 (18.3) | 549 (33.4) | 249 (33.2) | 225 (41.7) | 139 (28.7) | <0.001 |
| Intracerebral Hemorrhage | --- | --- | --- | --- | --- | 0.243 |
| Liver Disease | --- | 53 (3.2) | 15 (2.0) | 15 (2.8) | --- | <0.001 |
| Obesity | --- | 176 (10.7) | 74 (9.9) | 38 (7.0) | 29 (6.0) | <0.001 |
| Obstructive Sleep Apnea (OSA) | --- | 54 (3.3) | 15 (2.0) | --- | --- | <0.001 |
| Stroke | --- | 24 (1.5) | --- | --- | --- | 0.054 |
| **Mortality within 7 days, n (%)** | 148 (24.2) | 118 (7.2) | 93 (12.4) | 124 (23.0) | 27 (5.6) | <0.001 |

Demographics (age, gender, ethnicity, race, and past medical history) for all patients (N = 4029) included in study. Inter-hospital comparisons for categorical data were assessed with chi-square tests and numerical data using Kruskal-Wallis tests with Bonferroni-adjusted p-values reported. Values with fewer than ten patients per field are not provided to protect patient privacy.

To promote replicability, we placed our study within the Transparent Reporting of a Multivariable Prediction Model for Individual Prognosis or Diagnosis (TRIPOD) guidelines (Supplementary Table 3) and released our code under the GPLv3 license (Supplementary Materials). Software information can be found within the Supplementary Materials.

### Definition of Outcomes

The primary outcome was mortality within seven days of admission.

### Model Development and Selection

We generated two baseline conventional predictive models - a multilayer perceptron (MLP) and logistic regression with L1-regularization, or least absolute shrinkage and selection operator (LASSO). For consistency and to enable direct comparisons, each MLP model was built with the same architecture. We provide more information on model architecture and tuning in the Supplemental Materials. MLP and LASSO models were fit on all five hospitals.

Our primary model of interest was a federated learning model. In this model, training was performed in different sites, and parameters were shared to a central location, as depicted in Figure 1C. The process was as follows: a central aggregator initialized the federated model with random parameters. This model was sent to each site, then trained for one epoch. Next, model parameters were sent back to the central aggregator where federated averaging was performed. Updated parameters from the central aggregator were then sent back to each site, and this cycle was repeated for multiple epochs. Federated averaging scales the parameters of each site according to the number of available data points and sums all parameters by layer. Through this technique, federated models did not receive any raw data.

### Experimental Evaluation

All models were trained and evaluated using 10-fold stratified cross validation. We utilized the models’ probability scores to calculate areas under the receiver operating characteristic curve (AUC-ROC) with averages across the 10 folds.

## RESULTS

### Intercohort Comparisons

EHR data consisted of patient demographics, past medical history (Table 1), and admission vitals and labs (Supplementary Table 1). We found significant differences across hospitals in the proportion of outcome, specifically mortality within seven days (p<0.001), which ranged from 5.6% to 24.2% (Table 1). There were also significant differences in gender (p=0.004), age (p<0.001), ethnicity (p<0.001), and race (p<0.001). We found significant differences in the majority of key clinical features such as past medical history like asthma (p<0.001) and relevant lab tests including white blood cell counts (p<0.001; Supplementary Table 1).

### Classifier Training and Performance

LASSO and MLP models trained on data from each of the five MSHS hospitals (sites) separately (local), combined dataset (pooled), and via a federated learning framework (federated). All three training strategies for both models were evaluated on all sites (Figure 1B, Figure 1C). Results for model optimization (Supplementary Figure 2) and class balancing experiments (Supplementary Table 2) can be found in the Supplementary Materials. Final model hyperparameters are listed in Supplementary Table 4.

### Learning Framework Comparisons

Performance of all LASSO and MLP models (local, pooled, federated) was assessed at each site (Table 2, Figure 3). Federated LASSO outperformed the site’s local LASSO at all sites except MSQ, with AUC-ROC ranging from 0.693 to 0.805. LASSO pooled outperformed federated LASSO at all five sites and had AUC-ROCs ranging from 0.736 to 0.822.

**Table 2:** Model Performance By Site.

|  |  | Mount Sinai<br>Brooklyn<br>(MSB) | Mount Sinai<br>Hospital<br>(MSH) | Mount Sinai<br>Morningside<br>(MSM) | Mount Sinai<br>Queens<br>(MSQ) | Mount<br>Sinai West<br>(MSW) |
| --- | --- | --- | --- | --- | --- | --- |
| **Patients** |  | 611 | 1644 | 749 | 540 | 485 |
| **LASSO** | Local | 0.800 (0.742 -<br>0.857) | 0.704 (0.643 -<br>0.764) | 0.676 (0.619 -<br>0.734) | 0.733 (0.680 -<br>0.786) | 0.454<br>(0.281 -<br>0.626) |
|  | Pooled | 0.816 (0.759 -<br>0.872) | 0.797 (0.762 -<br>0.831) | 0.791 (0.746 -<br>0.835) | 0.736 (0.693 -<br>0.779) | 0.822<br>(0.735 -<br>0.909) |
|  | Federated | 0.802 (0.747 -<br>0.857) | 0.773 (0.740 -<br>0.807) | 0.776 (0.725 -<br>0.828) | 0.693 (0.643 -<br>0.744) | 0.805<br>(0.726 -<br>0.885) |
| **MLP** | Local | 0.833 (0.792 -<br>0.873) | 0.762 (0.710 -<br>0.814) | 0.739 (0.679 -<br>0.799) | 0.795 (0.756 -<br>0.834) | 0.739<br>(0.613 -<br>0.864) |
|  | Pooled | 0.825 (0.774 -<br>0.876) | 0.761 (0.687 -<br>0.835) | 0.726 (0.671 -<br>0.782) | 0.754 (0.721 -<br>0.788) | 0.829<br>(0.742 -<br>0.916) |
|  | Federated<br>No Noise | 0.827 (0.771 -<br>0.883) | 0.801 (0.743 -<br>0.858) | 0.796 (0.733 -<br>0.858) | 0.822 (0.786 -<br>0.858) | 0.834<br>(0.742 -<br>0.925) |
|  | Federated<br>With<br>Noise | 0.812 (0.754 -<br>0.869) | 0.767 (0.710 -<br>0.825) | 0.785 (0.724 -<br>0.846) | 0.822 (0.789 -<br>0.855) | 0.830<br>(0.743 -<br>0.917) |

**Figure 2.**
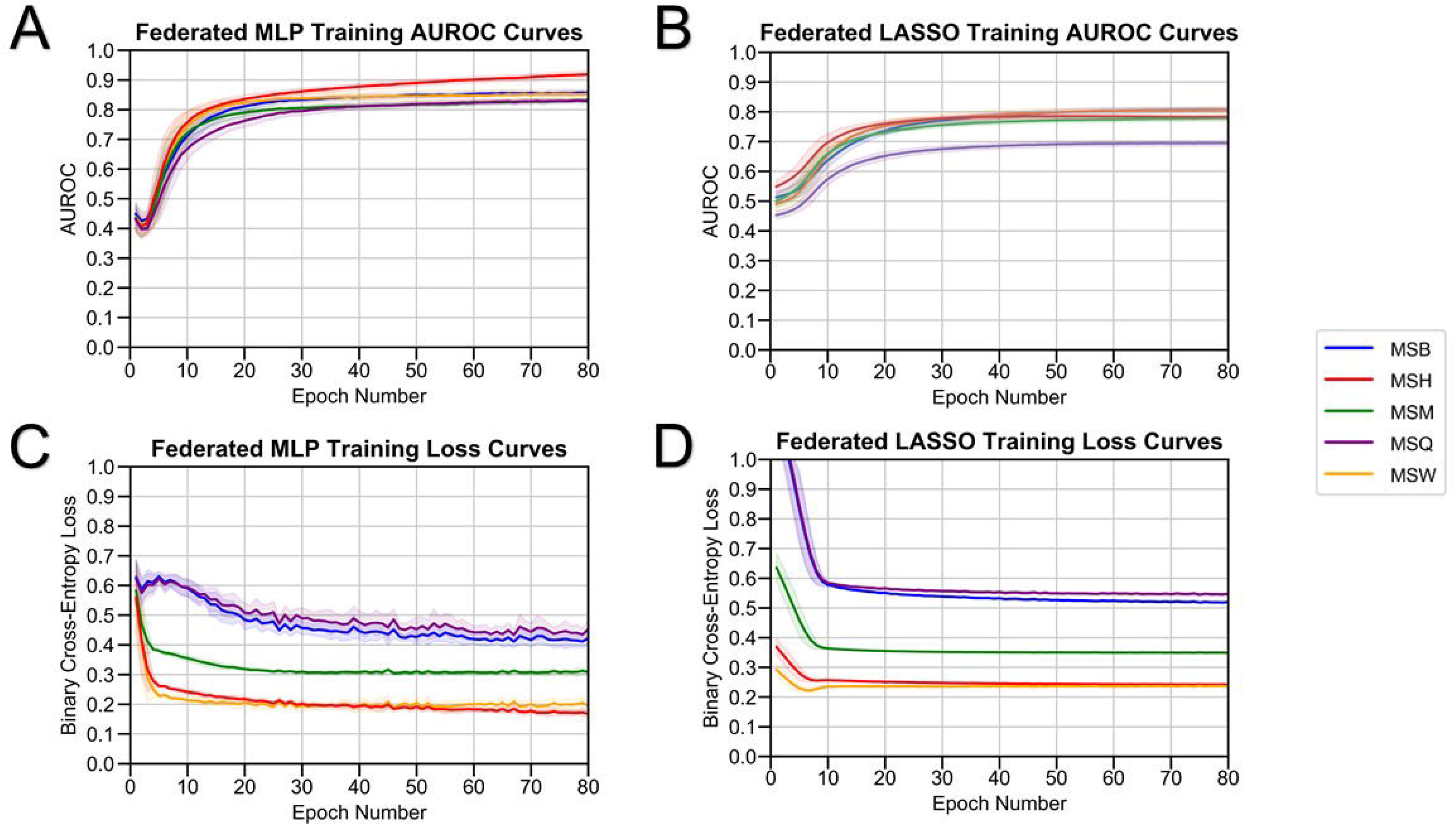
Federated Model Training. Performance of (A) federated MLP and (B) federated LASSO models as measured by area under the receiver-operating characteristic (AUC-ROC) versus the number of training epochs. Binary Cross-Entropy Loss of (C) Federated MLP and (D) Federated LASSO versus the number of training epochs.

**Figure 3.**
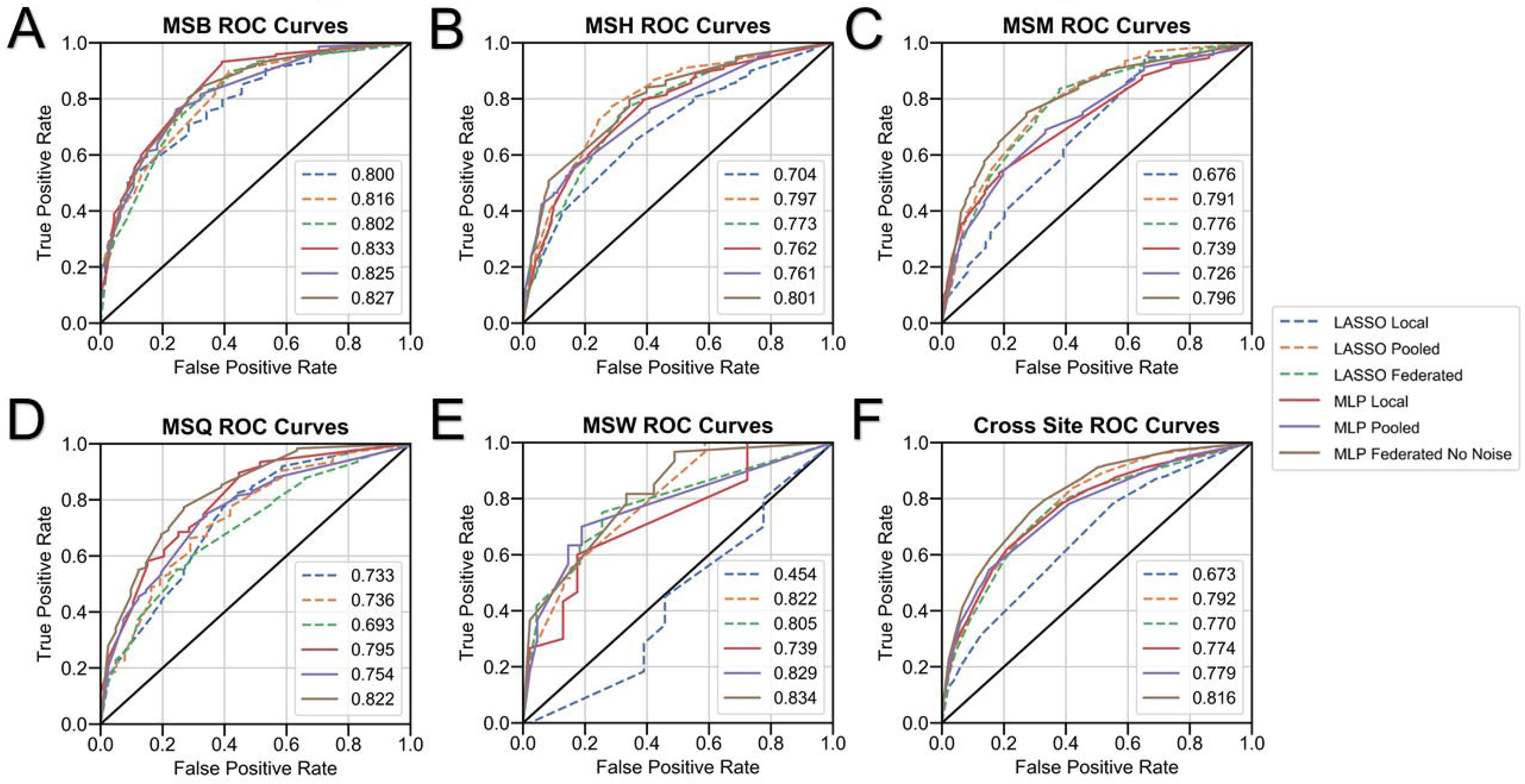
Model Performance by Site. Performance of all models (LASSO local, LASSO pooled, LASSO federated, MLP local, MLP pooled, MLP federated (no noise) by area under the receiver-operating characteristic (AUC- ROC) at (A) Mount Sinai Brooklyn (MSB) (n=611) (B) Mount Sinai West (MSW) (n=485), (C) Mount Sinai Morningside (MSM) (n=749), (D) Mount Sinai Hospital (MSH) (n=1644), and (E) Mount Sinai Queens (MSQ) (n=540). Averages of receiver-operating characteristic after 10-fold cross validation are shown. Average performance of each model across all five sites is presented in (F).

Performance of local, pooled, and federated LASSO and MLP models at each site as measured by area under the receiver-operating characteristic (AUC-ROC) with 95% confidence intervals. MLP pooled performed worse than local MLP at all hospitals except MSW, with MLP pooled AUC-ROCs varying from 0.754 to 0.829 while local MLP achieved AUC-ROCs ranging from 0.739 to 0.833. Federated MLP performed better than local MLP at all hospitals except MSB and performed better than the MLP pooled at all sites.

### Cross-model Comparisons

Local MLP outperformed local LASSO at all hospitals. Federated MLP surpassed federated LASSO at all sites. While performance varied slightly among both classifiers in different experiments, federated MLP consistently outperformed all other tested models at all hospitals except MSB. Additional performance metrics (AUPRC, accuracy, sensitivity, specificity, and F1- score) are described in Supplementary Table 5.

### Effect of Gaussian Noise

Gaussian noise introduced into federated MLP led to decreased performance at all sites except MSQ, where both had AUC-ROCs of 0.822. AUC-ROCs ranged from 0.796 to 0.834 in federated MLP without noise and 0.767-0.830 in federated MLP with noise (Supplemental Figure 1).

## DISCUSSION

To our knowledge, this is the first study to evaluate the efficacy of applying federated learning to predict mortality for COVID-19 using clinical data from EHRs. The data from the five hospitals used in this study represent a demonstrative use-case for this experiment. With disparate patient characteristics per hospital in terms of demographics, outcomes, sample size, and lab values, this study reflects a real-world scenario where federated learning can be leveraged for hospitals with diverse patient populations.

The primary findings of this study demonstrate that federated MLP and LASSO models outperformed their respective local models at most sites. Federated LASSO did not outperform pooled LASSO at any hospital. Federated MLP outperformed local MLP and federated LASSO at all hospitals. Federated MLP also outperformed pooled MLP at all sites which might be due to the experimental condition where the same underlying architecture was used for all MLP models. While this framework allowed for consistency in comparisons across learning strategies, it may have led to improper tuning of the pooled models. Collectively, these results exhibit the potential of federated learning to overcome drawbacks of fragmented local models.

Future inclusion of more sites into the federated learning network could continue to bridge the gap between pooled models, especially for LASSO.

Our results illustrate scenarios in which federated models should either be approached with caution or favored. MSQ was the only hospital where federated LASSO performed slightly worse than the local model with a difference of 0.04 in AUC-ROC, which may be attributed to a low sample size of 540 patients and a high mortality prevalence of 23.0% compared to other sites. However, at MSW, local LASSO severely underperformed in comparison to federated LASSO, with a difference of 0.351. MSW had the lowest sample size of all hospitals at 485 and the lowest COVID-19 mortality prevalence of 5.6%. This pattern emphasizes the benefit of federated learning for sites with small sample sizes and large class imbalance.

We also assessed the impact of adding noise to federated MLP. Training metrics were similar to federated MLP without noise insertion (Supplemental Figure 2). An average difference of 0.013 in AUC-ROC across all hospitals in these two models demonstrates that noise may be inserted to increase data security without severely compromising federated model performance.

We note a few limitations of our study. First, data collection was limited to MSHS hospitals in NYC. This may limit model generalizability to hospitals in other regions. Also, this study focused on applying federated learning to predict outcomes based on patient EHR data in principle rather than creating an operational framework for immediate deployment. As such, there are various aspects of the federated learning process that this work does not address such as load balancing, convergence, and scaling. These models included only clinical data and could be enhanced by incorporating other modalities such as imaging or free-text. We only implemented two widely used classifiers within this framework, but other algorithms may perform better.

Finally, identical MLP architectures were used across all learning strategies for direct comparisons but could have been further optimized.

Future work will focus on accessibility and expanding analysis of federated models. We plan to release code written within common data model EHR formats to better facilitate scalability. We will study salient features of importance for federated models and analyze changes as data are added. Finally, we will integrate additional data types such as images to improve model performance. We aim to use this federated learning framework to predict other adverse outcomes in hospitalized COVID-19 patients such as acute kidney injury.

## CONCLUSION

Federated learning is an effective method to share ML models without jeopardizing patient privacy. It is invaluable in the context of COVID-19, where limited data are segregated across various institutions. We demonstrated that federated learning models outperformed locally trained models to predict mortality. Federated learning offers a promising opportunity to construct more robust predictive models that can be leveraged in this pandemic.

## Data Availability

This article is written following the TRIPOD (Transparent Reporting of a Multivariable Prediction Model for Individual Prognosis or Diagnosis) guidelines, which are further elaborated in Supplementary Table 3. Furthermore, we release all code used for building the classifier under the GPLv3 license in a public GitHub repository.

https://github.com/HPIMS/CovidFederatedMortality

## CONTRIBUTORS

BSG, FW, and GSN conceived, designed, and supervised the study. AV collected the data and AV, SKJ, and JX were involved in data analysis. AV and SKJ were involved in interpreting the results. AV, SKJ, JX, ST, AK, SL, SS, IP, JKDF, TW, KPW, MB, EK, FK, AC, SZ, RM, AWC, EB, ZAF, FW and BSG drafted the initial manuscript. All authors approved provided critical comments, edited, and approved of the manuscript in its final form for submission.

## ACKNOWLEDGMENTS

We thank the Clinical Data Science and Mount Sinai Data Warehouse teams for providing data. We appreciate all the nurses, physicians, and providers who contributed to the care of these patients. This work is for patients and their family members who were affected by this pandemic.

## ETHICS APPROVAL

This study has been approved by the Institutional Review Board at the Icahn School of Medicine at Mount Sinai (IRB-20-03271).

